# Impact of rural trauma team development on prehospital time, referral decision to discharge interval, and outcomes of neurological and musculoskeletal injuries: a cluster randomized controlled trial

**DOI:** 10.1101/2025.03.04.25323372

**Authors:** Herman Lule, Micheal Mugerwa, Anne Abio, Benson Oguttu, Andrew Kakeeto, Fiona Walsh, Harvé Monka Lekuya, Robinson Ssebuufu, Patrick Kyamanywa, Till Bärnighausen, Jussi P. Posti, Michael Lowery Wilson

## Abstract

**Background:** We assessed the impact of rural trauma team development course (RTTDC) on clinical processes and patient outcomes of motorcycle-accident-related neurological and/or musculoskeletal injuries in selected Ugandan hospitals.

**Methods:** Trial design: Two-arm, parallel, multi-period, cluster-randomized controlled trial.

Participants: Trauma care frontliners, and patients aged 2-80 years at three intervention and three control Ugandan hospitals (1:1 allocation).

Randomization: Hospitals were randomly allocated to intervention or control using permuted block sequences.

Blinding: Patient-participants and outcome assessors were blind to allocation. Intervention arm: 500 trauma care frontliners received RTTDC; patients received standard care.

Control arm: Patients received standard care without RTTDC for staff.

Primary outcomes: Time from accident to admission and from referral to discharge.

Secondary outcomes: 90-day mortality and morbidity related to neurological and/or musculoskeletal injuries.

We followed the CONSORT guidelines for cluster randomized trials.

**Results:** We analyzed 1003 participants (501 intervention, 502 control). The intervention arm had a shorter median prehospital time (1hr; IQR=0·50-2·00) and referral to discharge interval (median 2hrs; IQR=1·25-2·75) vs. [(2hrs; IQR=1·50-4·00) mean difference 1·13hrs, p<0·0001 and (4hrs; IQR=2·50-4·10), mean difference 1·39hrs, p<0·0001 in the control], respectively. The 90-day mortality was more than halved in the intervention (5%, 24/457) vs. (13%, 58/430) in the control arm (p<0·0001). Fewer participants in the intervention group had unfavorable Glasgow Outcome Scale scores (9%, 42/457) vs. (20%, 87/430), p<0·0001. No difference was found in musculoskeletal injury morbidity outcomes (p=0·567).

**Interpretation:** Rural trauma team development training improved organizational time efficiency and clinical outcomes for neurological injuries without negatively impacting musculoskeletal injury morbidity outcomes.

**Funding:** University of Turku Graduate School, Neurocenter-Turku University Hospital, Turku University Hospital Foundation, Center for Health Equity in Surgery and Anesthesia-University of California San Francisco.

**Ethical approval:** Uganda National Council for Science and Technology (Ref: SS 5082).

**Trial registration:** Pan African Clinical Trial Registry (PACTR202308851460352).

**Research in context:** *Evidence before this study:* Understaffing and injury-related mortality are highest in low-and middle-income countries (LMICs) where trauma education and training opportunities are scarce. Observational studies in the US suggest that rural trauma team development training can improve clinician knowledge and reduce pre-hospital intervals. However, there are no prospective, randomized controlled trials that have assessed the translation of this knowledge and training into clinical practice affecting patient outcomes in LMICs.

*Added value of this study:* In a multi-center, cluster-randomized controlled trial, we investigated the impact of rural trauma team development, training and coordination on pre- and intra-hospital intervals, and outcomes for neurological and musculoskeletal injuries in an African low-resource setting. Results showed a reduction in prehospital time and all-cause mortality by more than half, without worsening patient-reported trauma morbidity.

*Implications of all the available evidence:* Prognostic level II evidence from this trial supports that locally contextualized, trainee-led rural trauma team development interventional programs are feasible and improve clinical processes and patient outcomes in LMICs.

## INTRODUCTION

Globally, access to essential surgical services remains restricted. In Africa, shortages of healthcare workers and their inequitable distribution further impede access to both emergency and elective surgical care. As a result, a substantial proportion of the population in Africa lacks access to these critical services.^1^

Optimizing human resources capacity, particularly for trauma care, is critical for strengthening health systems, alongside infrastructure development.^2^ The provision of trauma education for non-specialist primary care providers remains a potential remedy, to address the shortage of healthcare human resources in Africa.^3^ While several training programs such as the advanced trauma life support (ATLS) and primary trauma course (PTC) have been introduced, financial barriers and limited assessment of the impact on patient outcomes have hindered the implementation of these education programs, particularly for frontline trauma care providers in low-and-middle-income countries (LMICs).^3,4^

Locally contextualized trauma education programs are needed in LMICs due to resource constraints limiting ATLS uptake.^5^ Other existing courses such as PTC face criticism for limited international recognition and impact, warranting further exploration of need-specific alternatives.^4^ Evidence from observational studies in USA suggests rural trauma team development course (RTTDC) improves hospital processes and clinician knowledge.^6,7,8^

The ability of the RTTDC model to improve each trauma team member’s performance makes it applicable in LMICs, where team and organizational efficiency are more desired to mitigate the understaffing. However, researchers have primarily evaluated the course’s impact on provider performance rather than patient outcomes as ethical constraints have precluded the evaluation of RTTDC intervention through individually randomized prospective clinical trials.^8^ Furthermore, the need to mitigate potential contamination between treatment arms necessitates a cluster randomization study design. Evaluation of surgical interventions outside of randomized controlled settings contributes to research implementation gaps, limiting their utility in clinical practice. Cumulative research findings from emerging contemporaneous studies are necessary to drive practice and policy changes.

While prior systematic reviews have identified improvements in clinician knowledge from trauma education programs in LMICs, the next critical step is to evaluate their impact on organizational efficiency and patient outcomes in the clinical setting.^3,4,5^ Therefore, in this cluster-randomized trial, we examined the impact of a locally contextualized rural trauma team development training (RTTDC) on hospital processes and clinical outcomes in an LMIC setting, focusing on time efficiency, mortality, and morbidity of motorcycle accident-related neurological and/or musculoskeletal injuries, previously identified as critical areas for strengthening rural trauma systems.^9^ This study defines rural trauma team development as the implementation of RTTDC and the establishment of trauma teams in rural Uganda, which were previously non-existent.

The trial reporting followed the CONSORT guidelines for cluster-randomized trials, and the exploratory ancillary analyses due to COVID-19 interruptions adhered to CONSERVE guidelines.^10,11^

### Null hypothesis

We based the specific objectives of this trial on the null hypothesis that RTTDC training has no effect on the clinical process efficiency and outcomes of neurological and musculoskeletal injuries at both the treatment arm and individual levels.

### Specific objectives

The study aimed to i) determine the effect of RTTDC training on crash to admission, and referral decision to hospital discharge intervals (primary outcomes); and ii) determine the effect of RTTDC training on all-cause 90-day mortality and morbidity of neurological and musculoskeletal injuries (secondary outcomes).

## METHODS

This was a pragmatic two-arm, parallel, multi-period, cluster-randomized trial conducted from August 2019 to August 2023, with a one-year of interruption from March 2020 to March 2021 due to the extenuating effects of the Covid-19 pandemic. Six (6) Ugandan rural regional referral hospitals (Level III trauma centers), including Jinja, Hoima, Fort Portal, Mubende, Kiryandongo, and Kampala International University Teaching Hospital were used. Together, these hospitals serve a total population of 11.1 million people in three cities and 39 rural districts. Following the national standard of care, all patients requiring referral were transferred to the one central Level I trauma center (Mulago National Referral Hospital), Uganda’s comprehensive resource for multidisciplinary tertiary trauma care and rehabilitation.

### Randomization and masking

We employed permuted block sequences using an open source software to randomize the hospitals (clusters).^12^ The software generated six random codes for cluster assignment and enrollment, which were determined and concealed by an offsite study administrator. Equal cluster sizes were assumed.

We randomly assigned clusters in a 1:1 allocation ratio either to an intervention arm (comprising three hospitals), where 500 trauma care frontliners received the fourth edition of the American College of Surgeons’ RTTDC training administered in a multi-period manner or to the control arm (also consisting of three hospitals), where trauma care frontliners did not receive the training.^6,13^ Both patients and outcome assessors (medical officers independent of the trial who prospectively enrolled individual patients) were blind to the allocation. Hospital administrators enrolled trauma care frontliners.

### Eligibility criteria

To be eligible to participate, a trauma center had to be a teaching hospital offering 24/7 emergency surgical care with access to blood banks and imaging (CT, X-ray, Ultrasound scans) as locally available or outsourced. Additionally, each hospital had to be staffed with surgery consultants, residents, interns, and medical students.

The participants in this trial were either trauma care frontliners or trauma patients. The eligible trauma care frontliners constituted rural trauma team networks and were either surgery residents, intern doctors, third-fifth year medical/allied health students, or road traffic police officers concerned with the emergency evacuation of injured patients. It was not permissible for trauma care frontliners to offer their services in a way that would cross the treatment arms. Eligible patient participants were those aged 2-80 years with motorcycle accident-related injuries presenting within 24 hours either at the intervention or control hospitals during the study period.

We excluded medical trainees with no previous exposure to surgery clinical rotation; pregnant women; mentally incapacitated individuals with no legally authorized representatives to provide informed consent; patients with documented stroke, passengers in a car at time of collision to control confounding for injury mechanism; and deaths before imaging or arrival at emergency departments. More detailed eligibility criteria are documented in the trial protocol.^14^

### Patient and public involvement

We assessed the feasibility of trial outcome tools through engagement with trauma care frontliners and patient care givers before study commencement. Further engagements were conducted by establishing rural trauma teams rooted at subcounty level and obtaining bidirectional feedback on trauma team performance through weekly audit meetings as detailed in the study protocols.^13,14^

### Sample size determination and power analysis

The published trial protocol details the power calculations and methodology.^14^ The total sample size of 1003 patient participants (501 intervention, 502 control) was estimated using an R shiny application, accounting for the clustering design effect, 80% power, 5% type I error, ICC of 0·02 (0·01-0·05), CAC of 0·8, equal cluster size allowing a coefficient of variation of 0·5, and 18% loss to follow-up.^15^ The expected mean difference in primary outcomes was 1·02 hours based on an observational study in USA.^16^ Interim analyses were performed by a blinded statistician on 28 August 2022 in which results showed no patient-reported adverse incidents directly attributable to the intervention which was the pre-set termination criteria.

### Outcome measures and methods of assessment

Detailed outcome measurements, analysis metrics, and assessment time points are illustrated in the published protocol.^14^ The pre-specified primary outcomes used measures of central tendency to compare between arms: (i) prehospital time (from the accident scene to arrival at the emergency department), and (ii) referral discharge interval (from time a referral decision is made to the time a patient exited the hospital gate) in hours as a measure of process improvement.

The secondary outcomes compared between arms: (i) proportions of all-cause 90-day mortality from the time of injury as a measure of physician-centered clinical outcomes, and (ii) morbidity of neurological and musculoskeletal injuries at 90-days from time of admission using Glasgow Outcome Scale (GOS) and Trauma Outcome Measure Scores (TOMS) respectively as a measure of patient-reported functional outcomes.^17,18^ Both GOS and TOMS were measured as final values as reported to outcome assessors, although their baseline equivalents, i.e., Glasgow Coma Scale (GCS) and Trauma Expectation Factor Score (TEFS) were captured to compare morbidity at baseline.^17,19^ GOS was stratified into unfavorable (score 1-3) versus favorable outcomes (score 4-5), whereas TOMS were dichotomized into unfavorable (TOMS < TEFS) vs. favorable (TOMS ≥ TEFS) in accordance with previous studies.^20,18^

These outcome measures were selected to capture the complex multidimensional effects of trauma from a diverse perspective, including physical, social, and psychological domains. Both the GCS and the GOS have consistently demonstrated excellent internal validity, reproducibility, and reliability in multi-center trials, moreover the tools had been validated for use in LMICs, with versions applicable in both adults and pediatric populations.^18^

Acknowledging the importance of end-user involvement in research, the study used patient-reported outcomes (10-item Likert TOMS scale reported as a percentage) to assess the level of pain, physical function, disability, satisfaction with treatment, and overall life satisfaction at 90-days, compared to baseline expectations at admission (10-item Likert TEFS scale). Negative domains such as pain, disability and activity cutdown were assigned a minus score, while positive domains relating to satisfaction with life post-injury received a positive score, with total scores ranging from -700 to 300. Since injuries are not mutually exclusive in clinical settings, both GCS/GOS and TOM/TEFS were administered for patients with both neurological and musculoskeletal trauma who were able to verbalize, and Kampala Trauma Score (KTS II) captured the number of serious injuries to control for confounding due to polytrauma. The KTS II was used for its validity and suitability in predicting mortality in LMICs compared to other trauma scores, in addition to its superiority of putting physiological and anatomical indicators such as age-related comorbidities and multiplicity of injuries into context.^21^

### Study variables

We captured asymptomatic and symptomatic traumatic brain injury (TBI) based on National Institute of Neurological Disorders case definition. We obtained head and brain CT images for participants who either met the New Orleans criteria or the Canadian CT head rule based on local applicable algorithms used at the study sites, whereas we used radiographs to evaluate limb fractures.^22^ Based on our baseline study, we obtained random variables earlier identified to influence injury outcomes in our settings including: sex, age, comorbidities, injury mechanism, road user category, helmet use, prehospital (transportation, first aid and time), referral decision intervals, head and brain CT results, definitive surgical treatment, multiplicity of injuries and injury severity based on GCS and KTS II.^23^ These variables align with the global surgery agenda to improve national surgical care plans in support of universal health coverage.

### Statistical analysis

We tested the normality of distribution and equality of variance using Shapiro-Wilks and Levene’s tests, respectively. For primary outcomes, a two-sample Wilcoxon rank-sum test was used to compare the median (interquartile range, IQR) due to the skewed nature of the data. For secondary outcomes, we used an adjusted Chi-square to compare the difference in proportions of all-cause 90-day mortality, and proportions of unfavorable GOS/TOMS. Where applicable, we reported the difference in means (95% Confidence Interval, CI) as a measure of precision. The median (IQR) time to event (injury to death) was compared using a two-sample Wilcoxon rank-sum test and visualized using Kaplan-Meier survival estimates. We used Hussey and Hughes’ extension of the fixed effects model to examine the variation of outcomes in treatment arms across multiple time periods, assuming no study interruptions.^9^

### Pre-planned subgroup analyses

We performed pre-planned subgroup analyses for sex, road user category, injury mechanisms, severity, and multiplicity of injuries. We utilized the “melogit” command for mixed effects logistic regression models, which permitted adjustment for confounding to examine factors associated with all-cause 90-day mortality and morbidity of neurological and musculoskeletal injuries. We used the mixed effects models to accommodate variations in baseline characteristics of random variables, intervention effects, and unbalanced data patterns that could have occurred between treatment groups resulting from loss to follow-up. These models assumed that missing data was random, thereby eliminating the need for multiple imputations (sensitivity analyses). The fixed effects variable was the treatment arm (intervention vs. control) as the unit of analysis with the odds ratios and their corresponding 95% CI as a direct estimate of the effect size. Intracluster correlation coefficients (ICC) were computed in the mixed effects restricted maximum likelihood (REML) regression model using the “estat icc” command as the ratio of between cluster variance of outcomes to total variance of outcomes between and within cluster.

### Additional analysis

We compared the baseline characteristics of participants who had missing endpoints due to loss to follow-up between the study arms. To account for potential confounding due to factors related to COVID-19 pandemic’s socioeconomic impact on emergency trauma services and livelihood, trial investigators conducted exploratory analyses comparing outcomes between participants recruited before and after the pandemic in accordance with the CONSERVE guidelines.^11^ We examined all covariates for confounding and effect modification using Cochran-Mantel-Haenszel statistics. All analyses and data visualization were performed in Stata 15·0. We considered a *p*-value <0·05 as statistically significant. Due to manuscript length constraints, ancillary studies detailing results of mixed effects regression analyses, barriers to injury care, and impact of rural trauma team training on providers’ knowledge which construed the tertiary outcomes of this trial are reported separately.^24,[preprints 25,26]^

### Ethical approval

The study adhered to ethical guidelines outlined by the Declaration of Helsinki, the 2014 Uganda National Council for Science and Technology (UNCST) guidelines and received ethical approval from the UNCST research and ethics committee (Ref: SS 5082). Written informed consent was obtained from all participants or their legal representatives prior to recruitment.

## RESULTS

The participant CONSORT flow diagram is summarized in (Figure 1). At the end of the study period, there were 1003 participants (501 intervention: 502 control), with an average cluster size of 167 in both groups (14 individuals per cluster per period).

**Figure 1:**
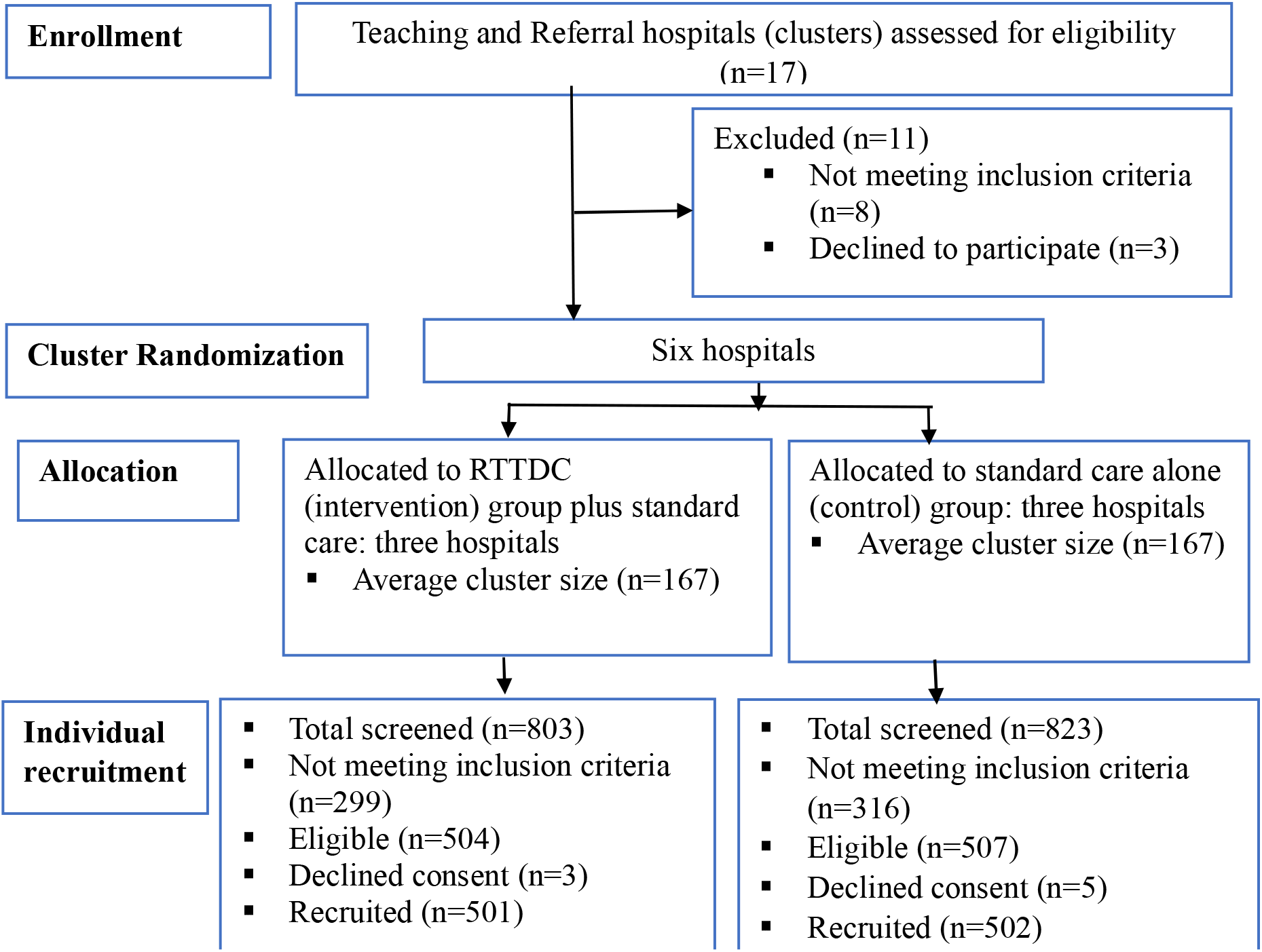
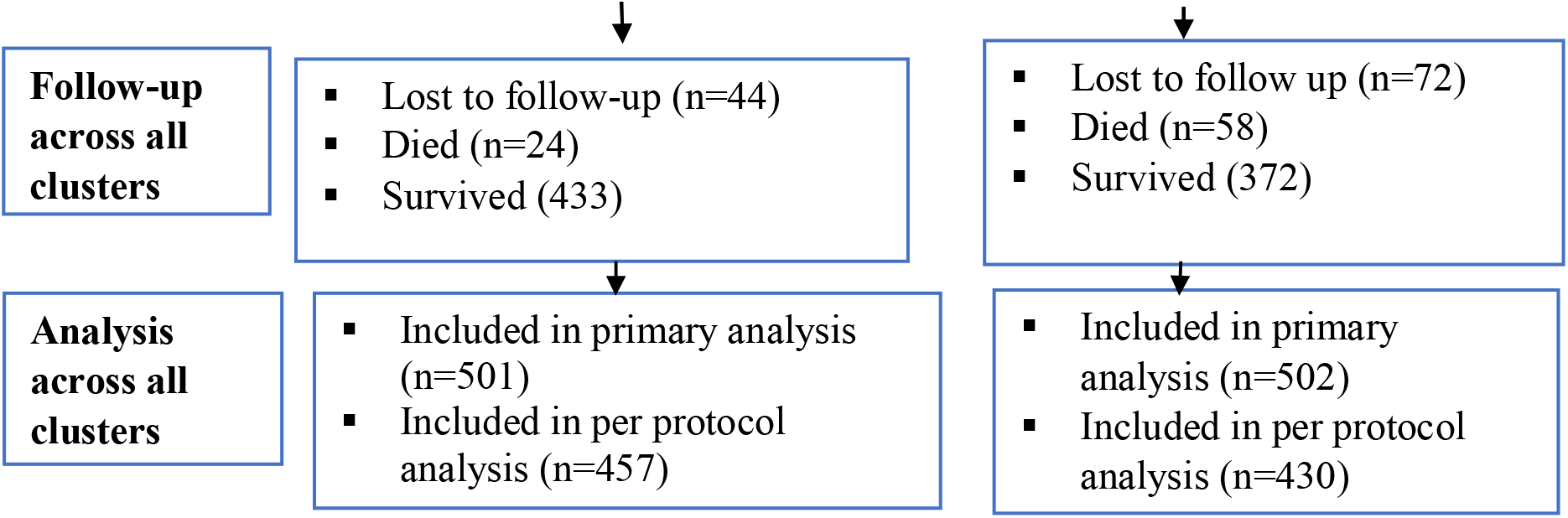
CONSORT flow diagram.

### Baseline sociodemographic and clinical characteristics

Cluster and individual level baseline sociodemographic and clinical characteristics are summarized in *Appendix 1*. Of 1003 participants, there were 82% (n=817) males vs. 19% (n=186) females [M:F (402:99) intervention vs. (415:87) control]. The overall median age (IQR) was 28 (22-37) years [28 (22-38) intervention vs. 28 (22-36) control]. There were no significant imbalances in the distribution of baseline characteristics between treatment groups except for 5 out of 37 covariates, i.e., employment status, proportions sustaining motorcycle-car crash, reported head impact, multiplicity of injuries, and respiratory rate. The overall median injury severity based on KTS II was 8, IQR (7-9), and did not differ significantly between groups (*p* = 0·360).

Irrespective of symptoms, 95% (949/1003) participants reported head trauma (impact). Symptomatic traumatic brain injury was present in 70% (699/1003) [69% (347/501) intervention vs. 70% (352/502) control]. We obtained head and brain CT images for 67% (675/1003) participants who met the CT criteria [67% (335/501) intervention vs. 68% (340/502) control]. The median GCS was 14, IQR (11-15), and did not differ between groups (*p* = 0·123). The admission systolic blood pressure was ≥ 90 mmHg in 82% (822), with oxygen saturation > 90% in 80% (802) of participants. Loss of consciousness and post-traumatic headache were the most reported clinical presentation. Acute epidural hematomas were the most observed radiological pathology (17.4%, 175/1003), followed by subdural hematomas (9.5%, 95/1003). Emergency craniotomy was the most performed neurosurgical intervention (23.5%, 236/1003). On the other hand, musculoskeletal injuries were present in 80% (799/1003), [80% (400/501) intervention vs. 80% (399/502) control] of which 27% (268/1003) had limb fractures [27% (134/501) intervention vs. 27% (134/502) control].

### Primary outcome I: comparison of prehospital time

For the 1003 participants, the overall median prehospital time was [2 hours, IQR (1·00-3·00)] and was shorter in the intervention compared to the control group [1 hour, IQR (0·50-2·00) vs. 2 hours, IQR (1·50-4·00)]. The mean difference was 1·13 hours, 95% CI: 0·96-1·29, *p* < 0·0001; [ICC = 0·265, 95% CI (0·022-0·854), *p* < 0·0001] (Figure 2). Adjusted predictions of prehospital time across study periods are presented in *Appendix 2*.

**Figure 2:**
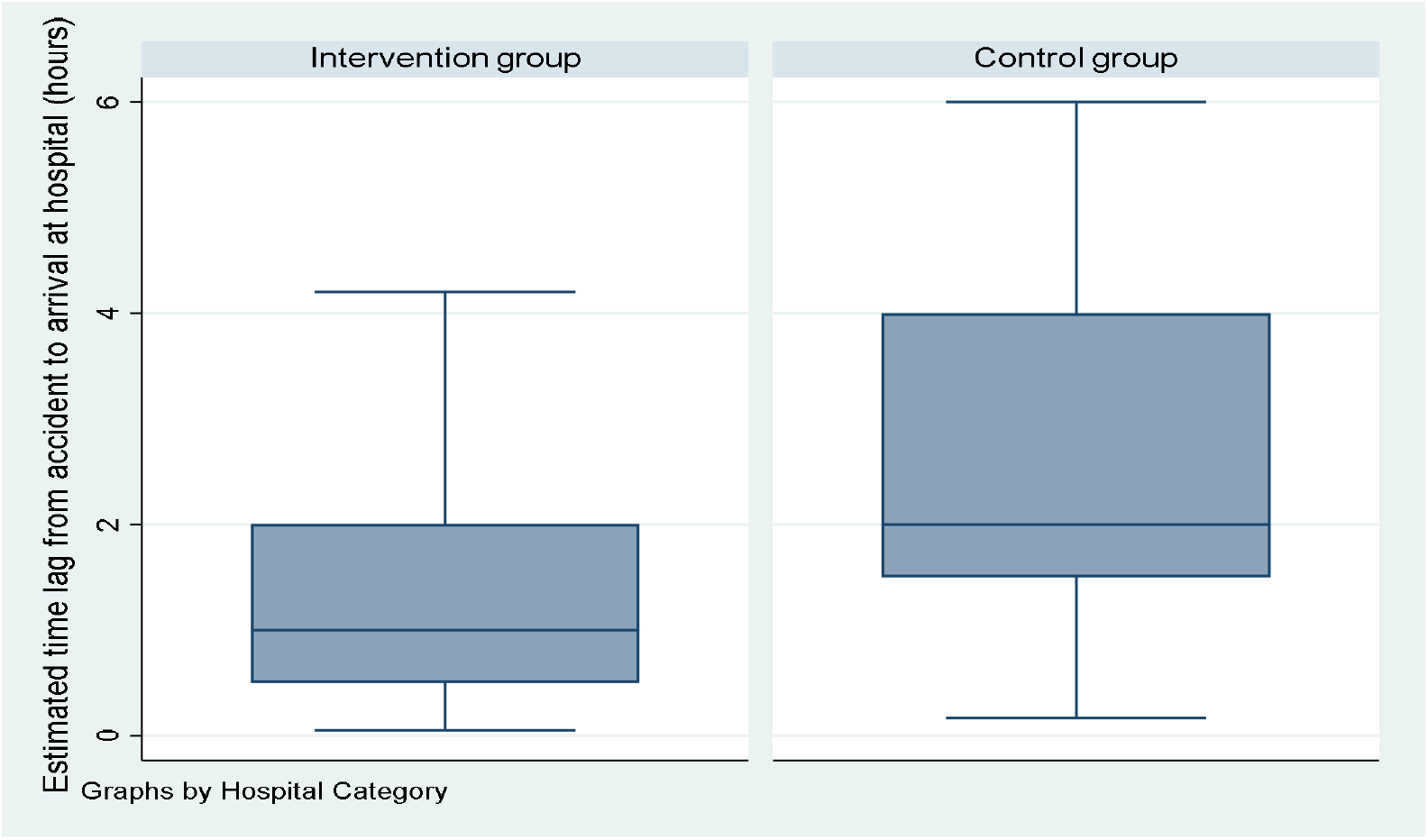
comparison of the median prehospital time lag from accident scene to arrival at emergency departments.

### Primary outcome II: comparison of referral decision to discharge interval

For the 691/1003 patients whose care demands exceeded local resources and warranted referral, the overall median referral decision to discharge interval was [3 hours, IQR (1·75-3·85)]. As presented in Figure 3, the referral decision to discharge interval was shorter in the intervention [2 hours, IQR (1·25-2·75)] vs. control group [4 hours, IQR (2·50-4·10)], with a mean difference of 1·39 hours, (95% CI: 1·23-1·55), *p* < 0·0001; [ICC = 0·444, 95% CI (0·047-0·928), *p* < 0·0001].

**Figure 3:**
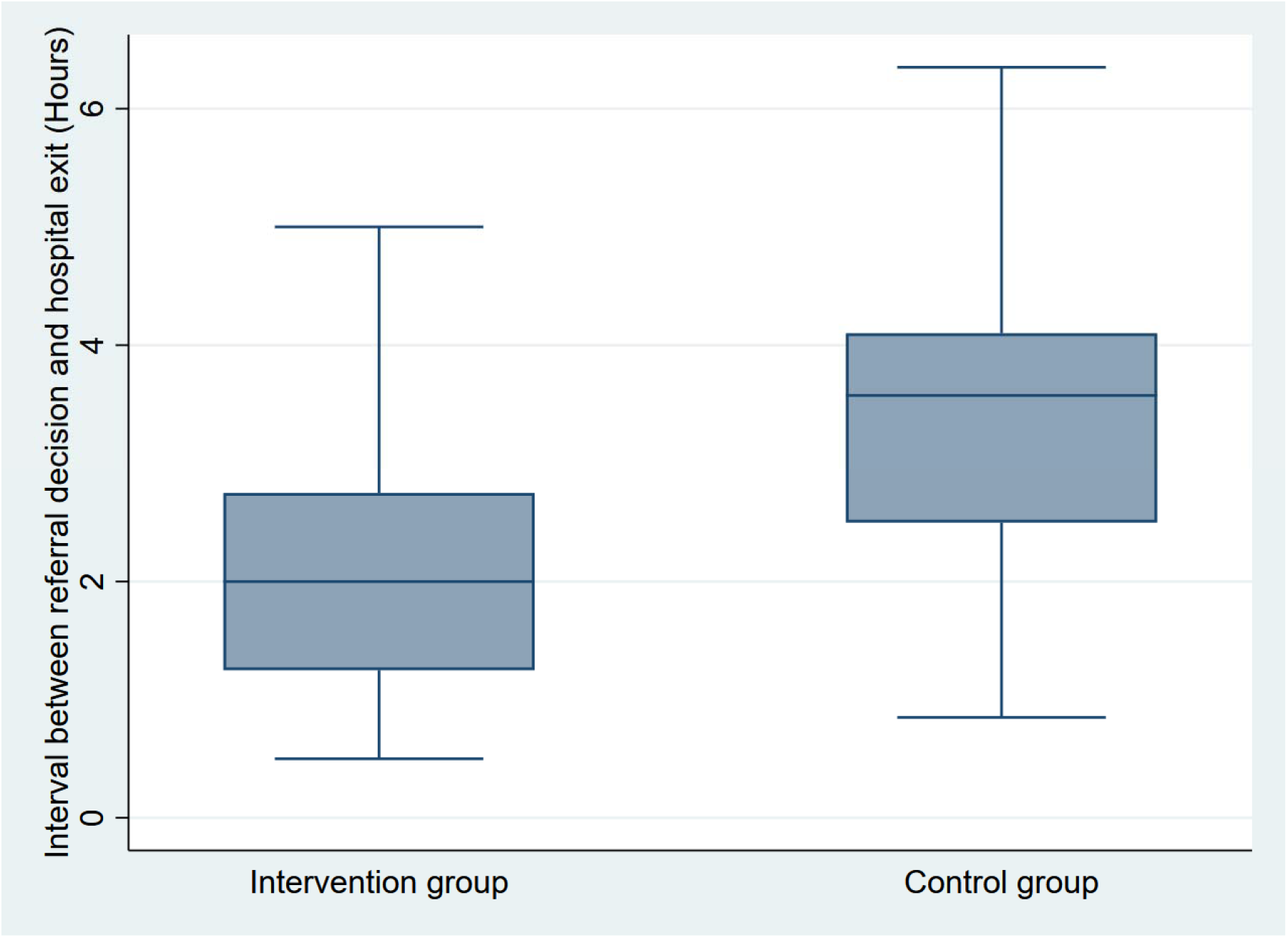
Comparison of referral decision to hospital discharge interval between groups.

#### Secondary outcome I: all-cause 90-day mortality

Of the 88% (887/1003) participants who completed 90-day follow-up, the overall 90-day mortality was 9% (82/887). The mortality was lower in the intervention at 5% (24/457) compared to the control group at 13% (58/430); [*X*^2^ (1, *N* = 887) =17·91, *p* < 0·0001]. Of the 82 deaths, eight occurred in the emergency departments of the control group versus zero in the intervention group. The overall median survival time amongst the remaining 74 mortalities was 3 days, IQR (1-8) and did not differ significantly between groups (*p* = 0·484) (Fig 4). Although more males (84%, n = 69) died than females (16%, n = 13), preplanned subgroup analyses showed that the median survival time did not differ significantly across study periods by sex (*p* = 0·707) (*Appendix 3*), and by road user category (*p* = 0·462) (*Appendix 4*).

**Figure 4:**
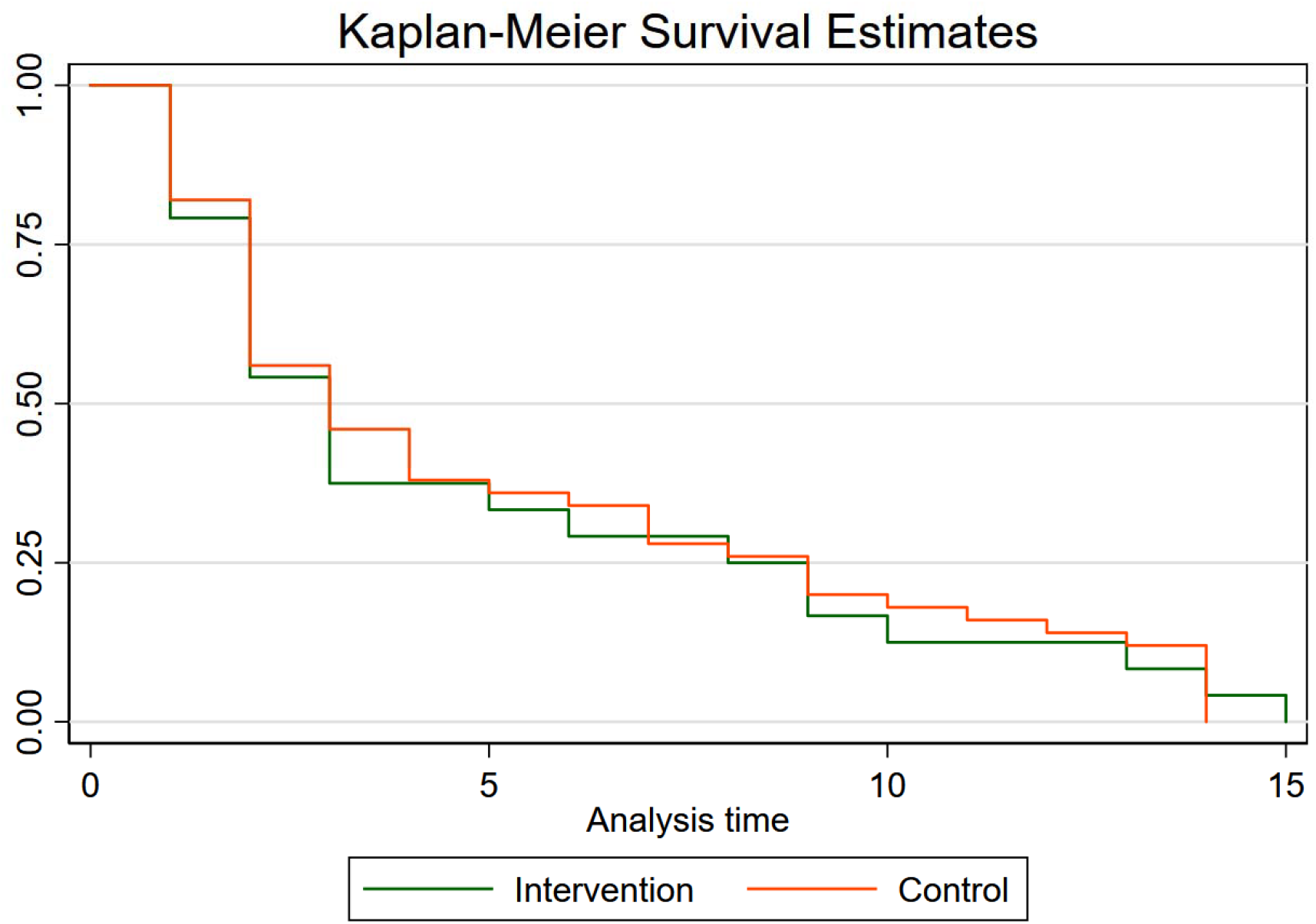
Comparison of Kaplan-Meier survival time in days between groups.

### Secondary outcome II: comparison of morbidity of neurological injuries

Of the 887/1003 participants who completed the 90-day follow-up, 86% (758/887) had a favorable Glasgow Outcome Scale (GOS 4-5). The proportion of participants with favorable outcomes was significantly higher in the intervention group (91%, 415/457) compared to the control group (80%, 343/430), [*X*^2^ (1, *N* = 887) =21·74, *p* < 0·0001]. The median GOS (IQR) was 5 (4-5) in both arms with the intervention group showing a slightly higher median compared to the control group (*p* = 0·003). The ICC was 0·037 (95% CI: 0·002-0·423, *p* < 0·0001). The results of all GOS subcategories amongst participants are summarized in *Appendix 5*.

### Secondary outcome III: comparison of morbidity of musculoskeletal injuries

Of the 637/799 patients with musculoskeletal injuries whose TEFS and TOMS were complete at 90-days, the majority (75%, 478/637) had a favorable outcome, i.e., (TOMS ≥ TEFS). The proportion of patients whose trauma outcome equaled or exceeded their initial expectations was comparable between the intervention 76% (262/345) vs. control 74% (216/292) [*X*^2^ (1, *N* = 637) =0.651, *p* = 0·567].

Assuming equal variance, the overall mean trauma expectation factor score (TEFS) was 80, 95% CI (69·2-91·3); [94, 95% CI (78·3-108·9) intervention vs. 67, 95% CI (50·5-82·5) control]. The mean difference was 27, 95% CI (5·0-49·2), *p* = 0·016; [ICC = 0·010, 95% CI (0·000-0·215), *p* = 0·041].

The overall mean trauma outcome measure score (TOMS) was 138, 95% CI (125·5-150·2); [145, 95% CI (128·4-161·7) intervention vs. 130, 95% CI (110·7-147·7) control]. The mean difference was 14, 95% CI (-9·0 to 40·9), *p* = 0·260; [ICC = 0·001, 95% CI (1·28e^-09^-0·997), *p* = 0·431] (Fig 5). The odds (OR) of unfavorable to favorable patient-centered trauma outcomes for musculoskeletal injuries did not differ significantly between intervention [OR 0·952, 95% CI (0·804-1·128) vs. control OR 1·073, 95% CI (0·907-1·269), *p* = 0·567]. This assertion remained valid even after preplanned subgroup analyses for limb fractures [OR 1·009, 95% CI (0·716-1·368) intervention vs. control OR 0·990, 95% CI (0·716-1·368), *p* = 0·950] and for isolated tibial fractures [OR 0·992, 95% CI (0·740-1·332) intervention vs. control OR 1·008, 95% CI (0·726-1·400), *p* = 0·960] (*Appendix 6*). Adjusted predictions of TEFS and TOMS across study periods are illustrated in *Appendix 7*.

**Figure 5:**
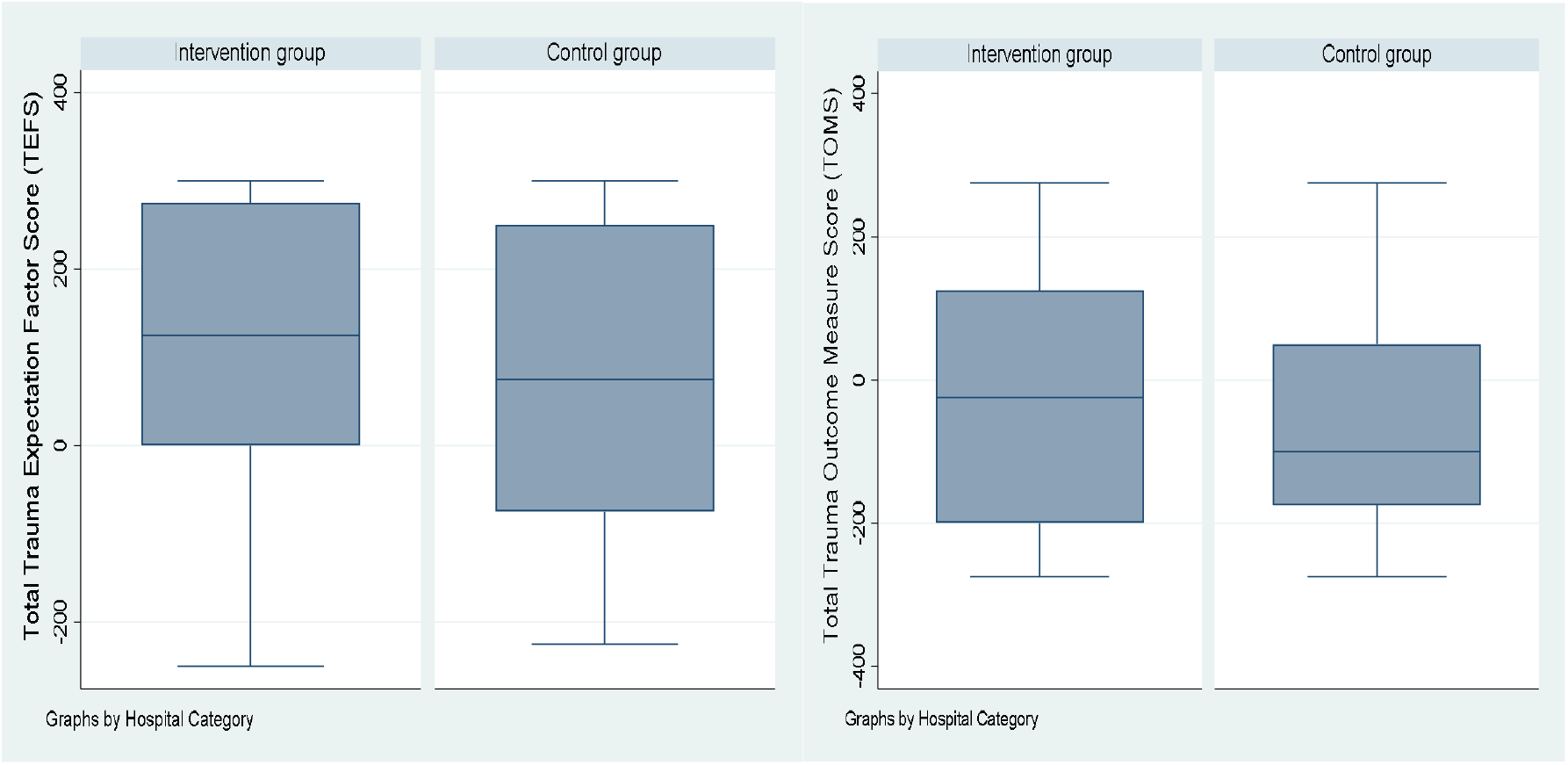
Comparison of trauma expectation factor scores (TEFS) and Trauma outcome measure scores (TOMS) between groups.

### Adverse events

There were no crossovers, but there were eight deaths at the emergency department in the control arm, which can be considered as an adverse injury outcome. At 90-day disposition amongst 805 retained survivors, a total of 16 (2%) participants were still hospitalized due to pressure ulcers sequelae to neurological injuries [(7/433) intervention vs. (9/372) control] whereas 789 were discharged home [(426/433) intervention vs. (363/372) control], p = 0·416. On evaluation of lost to follow-up (12%, n=116) who had missing secondary outcomes (90-day mortality, GOS, TOMS), the proportion was lower in the intervention 9% (44/116) vs. control 14% (72/116), *p* = 0·001, but the baseline sociodemographic and clinical characteristics of those lost to follow-up did not differ significantly between the two treatment groups (*Appendix 8*). The primary outcomes with missing end points were less than 0·01%. Our data quality and handling protocols in REDCap, have been previously described.^14^

To examine the extenuating circumstances of COVID-19, we performed exploratory analyses to probe any differences in outcomes and our results showed no differences in the prehospital interval (*p* = 0·038), referral decision-discharge interval (*p* = 0·588), and all-cause mortality (*p* = 0·434) before and after the COVID-19 pandemic. However, participants who were seen after the onset of the pandemic were twice as likely to report trauma outcome measure scores (TOMS) below their baseline expectations [OR 2·019; 95% CI (1·178 - 3·460), *p* = 0·011] and this association remained significant even after adjusting for confounding due to age, employment status, marital status, injury mechanisms, injury severity and multiplicity of injuries [aOR 2·095, 95% CI (1·199 – 3·659), *p* = 0·009].

## DISCUSSION

To our knowledge, this study represents a pioneering cluster-randomized trial assessing the impact of RTTDC on clinical processes and patient outcomes in an LMIC setting. This trial found that RTTDC training significantly reduced prehospital time by 1⍰13 hours, highlighting a potential for improved budget allocation for prehospital logistical systems aimed at enhancing care for neurological and orthopedic injuries in these settings. Prior research, primarily observational or quasi-experimental, has indicated similar reductions in referral decision times and transfer intervals, for instance a 1·91-hour decrease was reported in Western Virginia hospitals compared to control groups.^27^ Further findings from this trial indicated a reduction of 1⍰39 hours in the interval from referral decision to hospital discharge (p < 0·001). These results align with a recent study involving 472 trauma patients in the USA, which documented a similar decrease in emergency department dwell time.^7^ However, these studies often faced limitations due to small sample sizes and potential biases. Poor interhospital coordination and referral systems were the most important contributing factors to delayed emergency surgical access in rural settings, alongside human, financial, and infrastructural resource challenges as this trial demonstrated that trauma patients still arrive by public means in LMICs.

The trial’s overall 90-day mortality rate of 9·2% was comparable to prior Ugandan studies but higher than rates reported in USA and European cohorts.^28,29^ Notably, the intervention group exhibited significantly lower mortality rates than the control group, suggesting that RTTDC may effectively minimize mortality associated with prolonged referral decision times. In this trial, referral decision to dispatch time more than one hour was associated with four-fold increased mortality after controlling for confounding due to injury mechanisms and severity [aOR 4·215 (95% CI, 1·802-9·858, *p* = 0·001)]. This finding is particularly relevant given that delays in care have been shown to exacerbate injury-related mortality in both LMICs and HICs.^28^ Consistent outcomes were observed in previous studies, including a notable 50% reduction in mortality rates associated with RTTDC training in USA.^16^ Despite these positive results, variations in mortality outcome rates across studies exist, and may stem from different methodological approaches, sample sizes, and settings.^7^

In addition to mortality, the study explored morbidity outcomes, particularly concerning neurological and musculoskeletal injuries. The intervention group had a lower proportion of unfavourable Glasgow Outcome Scale (GOS) scores compared to controls, indicating improved recovery trajectories. However, the analysis of trauma outcome measures (TOMS) for musculoskeletal injuries was inconclusive, possibly due to higher proportions of polytrauma cases [OR 1·762, 95% CI (1·001 – 3·100, *p* = 0·049] and higher initial trauma outcome expectations among intervention participants. Existing literature corroborates the increased morbidity associated with polytrauma, emphasizing the need for future research into the long-term outcomes of RTTDC interventions in the context of multiple injuries.^30^

The implications of these findings are threefold. First, while RTTDC training shows promise for enhancing clinical processes and patient outcomes in LMICs, local and regional evaluations of its impact are essential for effective implementation. Establishing regional certification centers for trauma care providers could facilitate this goal. Second, the development of context-specific rural trauma transfer guidelines is critical, incorporating local norms and logistical realities. This may necessitate broader participation from various healthcare professionals, including medical trainees and lay traffic police who provide immediate crash response, to strengthen rural trauma systems. Lastly, a tailored needs assessment for each LMIC is crucial to adapt trauma education programs, ensuring they address specific cultural and systemic challenges while promoting sustainability. Existing trauma education programs in LMICs lacked impact evaluation, cohesive local contextualization and common outcome metrics for reporting.^3^ This trial provided critical clinical processes and patient-reported indices that could constitute a core outcome set.

### Study strengths and limitations

This trial offers significant strengths in evaluating a locally contextualized trauma education program in resource-limited rural settings. It effectively demonstrates that training trauma care frontliners such as voluntary traffic police officers and medical trainees can enhance clinical practices and patient outcomes. We previously highlighted the impact of rural trauma team training on care providers’ knowledge, and this trial builds on that foundation by showing the translation of acquired knowledge into improved clinical processes.^24^ Early engagement with frontliners ensured that the trial was relevant, and the findings may inform future core outcome sets for similar trials in LMICs.

However, the trial has notable limitations. First, a higher loss to follow-up in the control group introduces potential systematic attrition bias, although the overall dropout rate (11⍰6%) remains within acceptable limits. Additionally, the Intracluster correlation coefficient (ICC) may not accurately represent variations across cluster sizes for secondary outcomes, a common issue in cluster-randomized trials. Secondly, some patients at intervention sites may not have received care from trained providers, limiting the intervention’s individual-level impact. Moreover, the generalizability of findings is constrained to those who underwent CT scans, excluding emerging prognostic factors for neurological injuries such as blood-based biomarkers. Thirdly, although exploratory analyses indicated no significant changes in prehospital time due to the COVID-19 pandemic, its broader social effects on patient-reported morbidity outcomes remained challenging to quantify. However, we are unaware of any other events that could have influenced referral decisions and morbidity outcomes. Additionally, the absence of pre-intervention data at the study sites represents a limitation. Ideally, if data had been available prior to training, a comparison of injury outcomes before and after the training would have been preferable to the current approach of making parallel comparisons. Lastly, we are aware that aggregating intracranial lesions as axial or extra-axial and dichotomizing the GOS and TOMS tools as favorable vs. unfavorable outcomes could have led to loss of information and left unmeasured residual confounding that can potentially contribute to some of the observed associations.

## Conclusions

This trial provided Level II evidence that rural trauma team development training significantly decreased prehospital time and referral decision intervals, potentially lowering motorcycle accident-related mortality from neurological and musculoskeletal injuries, without negatively impacting patient-reported morbidity. These findings highlight the importance of enhancing rural trauma team capabilities and support ongoing investment in trauma education programs. Future research should focus on large, multi-country studies using long-term trauma registries and patient-centered quality-of-life outcome measures tailored to local contexts in LMICs.

## Contributors

HL was the lead researcher who conceived, designed, and implemented the trial, and wrote the initial draft of the manuscript. BO, AK, HML, RS, and PK assisted with administrative tasks, data acquisition, participant follow-up, and critically reviewed the manuscript. HL, MM, and AA analyzed and cross-validated the data. TB critiqued the study methods. FW and TB critically reviewed the manuscript. HL, TB, JPP, and MLW sourced funding. JPP and MLW supervised the study and critically reviewed the manuscript. All authors read and approved the final manuscript.

## Reflexivity statement

A detailed statement regarding how partnerships and collaborations were actualized is available in *Appendix 9*.

## Acknowledgements

We are grateful to Drs. Asiimwe Daniel, Namutosi Esther, Kabarokole Annet, Mwine David, Ssekabembe John, Japheth Comfort, Nalumansi Hilda, and Rwamukaga Nasurudin for their invaluable assistance in the data collection process and for consenting to the publication of their names. Additionally, we thank Ms. Mary Nnabagulanyi for her support in the offsite administration and coordination of this trial and appreciate our research participants for their commitment and time dedicated to this study.

## Patient consent for publication

Not applicable.

## Ethics Approval

This trial was approved by the Research and Ethics Committees of Mbarara University of Science and Technology (Ref: MUREC 1/7; 05/5-19) and Uganda National Council for Science and Technology (Ref: SS 5082).

## Provenance and peer review

Not commissioned, pending peer review.

## Data availability statement

Deidentified participant data and analysis codes that underlie the findings of this study are available within this article as supplemental materials and within an open access data repository.^31^ Further details can be obtained from the study protocol and trial registry.^14^ https://pactr.samrc.ac.za/TrialDisplay.aspx?TrialID=25763

## Supplemental material

See appendices (1-9).

