## Appendices for "Impact of rural trauma team development on prehospital time, referral decision to discharge interval, and outcomes of neurological and musculoskeletal injuries: a cluster randomized controlled trial": CONSERVE-Checklists.docx

| CONSERVE-CONSORT Extension: Checklist^⸸^ | | | | | |
| --- | --- | --- | --- | --- | --- |
| Item | Item Title | Description | | | Page No. |
| I. | Extenuating Circumstances | Describe the circumstances and how they constitute extenuating circumstances. | | | 5, 8 |
| II. | Important Modifications | a. Describe how the modifications are important modifications. | | | 8, 13, 14 |
|  |  | b. Describe the impacts and mitigating strategies, including their rationale and implications for the trial. | | | 8, 13, 14 |
|  |  | c. Provide a modification timeline. | | | 5 |
| III. | Responsible Parties | State who planned, reviewed and approved the modifications. | | | 8, 14 |
| IV. | Interim data | If modifications were informed by trial data, describe how the interim data were used, including whether they were examined by study group, and whether the individuals reviewing the data were blinded to the treatment allocation. | | | 6 |
| CONSORT Number and Item | | For each row, if important modifications occurred check “direct impact” and/or “mitigating strategy” and describe the changes in the trial manuscript or supplement. Check “no change” for items that are unaffected in the extenuating circumstance. | | | Page No. |
|  |  | No Change | Impact* | Mitigating Strategy** |  |
| 1 | Title and abstract | X |  |  | N/A |
| 2 | Introduction | X |  |  | N/A |
| 3 | Methods: Trial Design | X |  |  | N/A |
| 4 | Methods: Participants | X |  |  | N/A |
| 5 | Methods: Interventions | X |  |  | N/A |
| 6 | Methods: Outcomes | X |  |  | N/A |
| 7 | Methods: Sample Size | X |  |  | N/A |
| 8-10 | Methods: Randomisation | X |  |  | N/A |
| 11 | Methods: Blinding | X |  |  | N/A |
| 12 | Methods: Statistical methods |  | X  (Potential confounding of study outcomes due to COVID-19 pandemic) | Additional exploratory analyses by stratification of data that were collected before and after COVID-19 pandemic | 8, 13, 14 |
| 13 | Results: Participant flow | X |  |  |  |
| 14 | Results: Recruitment |  | X (Inaccessibility to study participants due to lock downs and work constraints amongst data collection personnel due to prioritisation of COVID-19 related emergencies) | Recruitment was temporarily suspended for one year during March 2020 to March 2021. Since the study had been approved for 4 years, this had minimal impact as preplanned active data collection period had been 36 months amounting to 12 quarters (periods) of 3-months per quarter. | 5, 15 |
| 15 | Results: Baseline data |  | X  (Need to know the COVID-19 status of study participants for safety of research personnel and the rest of study participants) | COVID-19 status was added as a comorbidity on the data capture tool although no such cases were identified.    The COVID-19 screening process was introduced as routine standard operating procedures for all patients arriving at accident and emergency departments at all study centers in accordance with local hospital protocols, government regulations and research ethics committee | Trial protocol (Ref. 14) |
| 16 | Results: Numbers analysed | x |  |  | N/A |
| 17 | Results: Outcomes and estimation |  | X (Confounding due to socioeconomic impacts of COVID-19 on study outcomes) | Additional analysed performed to explore the confounding regarding how COVID-19 pandemic impacted the primary and secondary outcomes, after stratification of the data collected before and after the pandemic | 8, 13, 14 |
| 18 | Results: Ancillary analyses |  | X (Confounding due to socioeconomic impacts of COVID-19 on study outcomes) | Additional exploratory analyses were performed to evaluate how the pandemic impacted patient reported outcomes, outside of the preplanned analyses given the known resource impact of COVID-19 on emergency care services and its socioeconomic impacts at individual level. | Appendix 1, Pages 13, 14, 15 |
| 19 | Results: Harms |  | X  (Effect of COVID-19 on safety of research participants and personnel) | Personal protective equipment were introduced after at all the study sites in accordance with the local government regulations and research ethics committee to protect the personnel involved in data collection. | Study Protocol (Ref. 14) |
| 20 | Discussion: Limitations |  | x  (Potential confounding to patient reported outcomes) | How the pandemic could have impacted patient reported trauma outcome measures regarding social aspects of their quality of life after trauma is included in the study limitations | Pg 15 |
| 21 | Discussion: Generalisability | x |  |  | N/A |
| 23 | Other information: Registration |  | X  (The drive of COVID-19 to research literature bias and trial registration) | Retrospective trial registration due to prioritisation of COVID-19 related studies by the handling trial registrars | Trial protocol (Ref. 14) |
| 24 | Other information: Protocol |  | x (Publication bias of COVID-19 pandemic response-related studies) | Late protocol publication due to the prioritisation of COVID-19 related studies by the target journals and the general lack of timely reviewers during COVID-19 pandemic period | Trial protocol (Ref. 14) |
| 25 | Other information: Funding | x |  |  | N/A |
| *Aspects of the trial that are directly affected or changed by the extenuating circumstance and are not under the control of investigators, sponsor or funder.  **Aspects of the trial that are modified by the study investigators, sponsor or funder to respond to the extenuating circumstance or manage the direct impacts on the trial. | | | | | |
| ^⸸^ Orkin, A. M., Gill, P. J., Ghersi, D., Campbell, L., Sugarman, J., Emsley, R., Steg, P. G., Weijer, C., Simes, J., Rombey, T., Williams, H. C., Wittes, J., Moher, D., Richards, D. P., Kasamon, Y., Getz, K., Hopewell, S., Dickersin, K., Wu, T., Ayala, A. P., … CONSERVE Group (2021). Guidelines for Reporting Trial Protocols and Completed Trials Modified Due to the COVID-19 Pandemic and Other Extenuating Circumstances: The CONSERVE 2021 Statement. *JAMA*, *326*(3), 257–265. <https://doi.org/10.1001/jama.2021.9941> | | | | | |
