## Appendices for "Impact of rural trauma team development on prehospital time, referral decision to discharge interval, and outcomes of neurological and musculoskeletal injuries: a cluster randomized controlled trial": CONSORT-2010-Checklist extension for cluster.docx

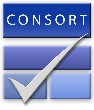
CONSORT 2010 statement checklist: extension to cluster randomised trial^1,2^

| Section/Topic | Item No | Checklist item | Reported on page No |
| --- | --- | --- | --- |
| Title and abstract | | | |
|  | 1a | Identification as a cluster randomised trial in the title | 1 |
|  | 1b | Structured summary of trial design, methods, results, and conclusions (for specific guidance see CONSORT for abstracts) | 2 |
| Introduction | | | |
| Background and objectives | 2a | Scientific background and explanation of rationale for using a cluster design | 4 |
|  | 2b | Specific objectives or hypotheses whether objectives pertain to cluster level, individual participant or both | 4,5 |
| Methods | | | |
| Trial design | 3a | Description of trial design (such as parallel, factorial) including allocation ratio | 5 |
|  | 3b | Important changes to methods after trial commencement (such as eligibility criteria), with reasons | 4,8 |
| Participants | 4a | Eligibility criteria for participants and for clusters | 5 |
|  | 4b | Settings and locations where the data were collected | 5 |
| Interventions | 5 | The interventions for each group with sufficient details to allow replication, including how and when they were actually administered (interventions pertain to the cluster level) | 5 |
| Outcomes | 6a | Completely defined pre-specified primary and secondary outcome measures, including how and when they were assessed (outcomes pertain to both cluster and individual levels) | 6 |
|  | 6b | Any changes to trial outcomes after the trial commenced, with reasons | 8 |
| Sample size | 7a | How sample size was determined including number of clusters assuming equal clusters, cluster size, coefficient of intracluster correlation (ICC or *k*) and an indication of its uncertainty | 6, protocol ref .14 |
|  | 7b | When applicable, explanation of any interim analyses and stopping guidelines | 6, protocol ref 14. |
| Randomisation: |  |  |  |
| Sequence generation | 8a | Method used to generate the random allocation sequence. | 5, protocol ref. 14 |
|  | 8b | Type of randomisation; details of any restriction (such as blocking and block size), details of stratification or matching if used | 5, protocol ref. 14 |
| Allocation concealment mechanism | 9 | Mechanism used to implement the random allocation sequence (such as sequentially numbered containers), describing any steps taken to conceal the sequence until interventions were assigned. Specification that allocation was based on clusters rather than individuals and whether allocation concealment (if any) was at the cluster level, the individual cluster level or both | 5, protocol ref. 14 |
| Implementation | 10a | Who generated the random allocation sequence, who enrolled clusters, and who assigned clusters to the interventions | 5 |
|  | 10b | Mechanism by which individual participants were included in the cluster for purposes of the trial (such as complete enumeration, random sampling) | 5 |
|  | 10c | From whom consent was sought (representatives of the cluster, or individual cluster members, or both, and whether consent was sought before or after the randomisation. | 8 |
| Blinding | 11a | If done, who was blinded after assignment to interventions (for example, participants, care providers, those assessing outcomes) and how | 5 |
|  | 11b | If relevant, description of the similarity of interventions | 5 |
| Statistical methods | 12a | Statistical methods used to compare groups for primary and secondary outcomes | 6-7 |
|  | 12b | Methods for additional analyses, such as subgroup analyses and adjusted analyses | 7-8 |
| Results | | | |
| Participant flow (a diagram is strongly recommended) | 13a | For each group, the number of clusters that were randomly assigned, received intended treatment, and were analysed for the primary outcome | Fig 1 |
|  | 13b | For each group, losses and exclusions for both clusters and individual cluster members | Fig 1 |
| Recruitment | 14a | Dates defining the periods of recruitment and follow-up | 5 |
|  | 14b | Why the trial ended or was stopped | 7,15 |
| Baseline data | 15 | Baseline characteristics for the individual and cluster levels as applicable for each group | Appendix 1 |
| Numbers analysed | 16 | For each group number of clusters included in each analysis | Fig 1 |
| Outcomes and estimation | 17a | Results at the individual or cluster level as applicable and a coefficient of intracluster correlation (ICC or *k*) for each primary outcome) | 9-12 |
|  | 17b | For binary outcomes, presentation of both absolute and relative effect sizes is recommended | Fig 2,3,5 |
| Ancillary analyses | 18 | Results of any other analyses performed, including subgroup analyses and adjusted analyses, distinguishing pre-specified from exploratory | Pg 11 & 13 Appendices 3,4,6,7,8 |
| Harms | 19 | All important harms or unintended effects in each group (for specific guidance see CONSORT for harms) | 13 |
| Discussion | | | |
| Limitations | 20 | Trial limitations, addressing sources of potential bias, imprecision, and, if relevant, multiplicity of analyses | 15 |
| Generalisability | 21 | Generalisability (external validity, applicability) of the trial findings to clusters and/or individual participants as relevant | 15 |
| Interpretation | 22 | Interpretation consistent with results, balancing benefits and harms, and considering other relevant evidence | 15 |
| Other information | | |  |
| Registration | 23 | Registration number and name of trial registry | 2 |
| Protocol | 24 | Where the full trial protocol can be accessed, if available | Ref.14 pg 17 |
| Funding | 25 | Sources of funding and other support (such as supply of drugs), role of funders | 2,16 |

1. Campbell MK, Piaggio G, Elbourne DR, Altman DG; CONSORT Group. Consort 2010 statement: extension to cluster randomised trials. BMJ. 2012;345:e5661. Published 2012 Sep 4. doi:10.1136/bmj.e5661
2. Schulz KF, Altman DG, Moher D; CONSORT Group. CONSORT 2010 statement: updated guidelines for reporting parallel group randomised trials. BMJ. 2010;340:c332. Published 2010 Mar 23. doi:10.1136/bmj.c332
