## Appendices for "Impact of rural trauma team development on prehospital time, referral decision to discharge interval, and outcomes of neurological and musculoskeletal injuries: a cluster randomized controlled trial": Supplementary Materials.docx

**Supplementary material**

[Appendix 9: Reflexivity statement regarding collaborative partnerships. [1]	11](#_Toc190221639)

### **Appendix 1: Baseline sociodemographic and clinical characteristics of participants.**

| **Characteristic** | **Categories** | **Intervention Group (N=501)**  n(%) | | **Control Group (N=502)**  n(%) | | **Overall (N=1003)**  n(%) | **p-value^a^** |
| --- | --- | --- | --- | --- | --- | --- | --- |
| General demographic and clinical characteristics across all participants | | | | | | | |
| Age (Years) | Median (IQR) | 28 (22.0-38.0) | | 28 (22.0-36.0) | | 28 (22.0-37.0) | .826^t^ |
| Sex | Male | 402 (80.2) | | 415 (82.7) | | 817 (81.5) | .322 |
|  | Female | 99 (19.8) | | 87 (17.3) | | 186 (18.5) |  |
| Marital status | Single | 184 (36.7) | | 197 (39.2) | | 381 (38.0) | .297 |
|  | Married | 286 (57.1) | | 285 (56.8) | | 571 (56.9) |  |
|  | Divorced | 21 (4.2) | | 16 (3.2) | | 37 (3.7) |  |
|  | Other | 10 (2.0) | | 4 (0.8) | | 14 (1.4) |  |
| Employment status | Formal paid employment | 33 (6.6)* | | 61 (12.2)* | | 94 (9.4) | .017* |
|  | Self-employed | 349 (69.7)* | | 319 (63.5)* | | 668 (66.6) |  |
|  | Unemployed | 25 (5.0) | | 36 (7.2) | | 61 (6.1) |  |
|  | Student | 83 (16.6) | | 76 (15.1) | | 159 (15.9) |  |
|  | Other | 11 (2.2) | | 10 (2.0) | | 21 (2.1) |  |
| Commute distance (Km) | Median (IQR) | 5.0 (2.0-10.0) | | 5.0 (2.0-10.0) | | 5.0 (2.0-10.0) | .065^t^ |
| Road user category | Passenger | 175 (34.9) | | 153 (30.5) | | 328 (32.7) | .317 |
|  | Pedestrian | 93 (18.6) | | 102 (20.3) | | 195 (19.4) |  |
|  | Motorcyclist | 233 (46.5) | | 247 (49.2) | | 480 (47.9) |  |
| Injury Mechanism | Motorcycle-motorcycle crash | 206 (41.1) | | 225 (44.8) | | 431 (43.0) | .028* |
|  | Motorcycle-pedestrian | 114 (22.7) | | 139 (27.7) | | 253 (25.2) |  |
|  | Motorcycle-car crash | 113 (22.6)* | | 87 (17.3)* | | 200 (19.9) |  |
|  | Motorcycle-static object | 68 (13.6) | | 51 (10.2) | | 119 (11.9) |  |
| Means of arrival | Ambulance | 96 (19.2) | | 103 (20.5) | | 199 (19.8) | .443 |
|  | Public means (taxi/motorcycle) | 398 (79.4) | | 378 (75.3) | | 776 (77.4) |  |
| Systolic blood pressure at admission (mmHg) | ≤ 49 | 0 (0.0) | | 2 (0.4) | | 2 (0.2) | .366 |
|  | 50 - 89 | 89 (17.8) | | 90 (17.9) | | 179 (17.8) |  |
|  | > 90 | 412 (82.2) | | 410 (81.7) | | 822 (82.0) |  |
| Respiratory rate at admission (breaths per minute) | ≤ 9 | 4 (0.8) | | 4 (0.8) | | 8 (0.8) | .020* |
|  | 10-29 | 388 (77.4)* | | 423 (84.3)* | | 811 (80.9) |  |
|  | ≥ 30 | 109 (21.8)* | | 75 (14.9)* | | 184 (18.3) |  |
| Oxygen circulation at admission | > 90% | 398 (79.4) | | 404 (80.5) | | 802 (80.0) | .682 |
|  | ≤ 90% | 103 (20.6) | | 98 (19.5) | | 201 (20.0) |  |
| Pre-hospital care (first aid) received before arrival) | No | 282 (56.3) | | 262 (52.2) | | 544 (54.2) | .193 |
|  | Yes | 219 (43.7) | | 240 (47.8) | | 459 (45.8) |  |
| Evidence of any chronic medical illness ^e^ | No | 463 (92.4) | | 477 (95.0) | | 940 (93.7) | .089 |
|  | Yes | 38 (7.6) | | 25 (5.0) | | 63 (6.3) |  |
| Multiplicity of serious injuries | > One | 127 (25.3)* | | 94 (18.7)* | | 221 (22.0) | .019* |
|  | One | 291 (58.1)* | | 332 (66.1)* | | 623 (62.1) |  |
|  | None | 83 (16.6) | | 76 (15.1) | | 159 (15.9) |  |
| Injury severity score based on Kampala Trauma Score | Median (IQR) | 8 (7-9) | | 8 (7-9) | | 8 (7-9) | .360^t^ |
| Categories of injury severity based on Kampala Trauma Score (KTS) | (9-10) | 233 (46.5) | | 249 (49.6) | | 482 (48.1) | .320 |
|  | (7-8) | 165 (32.9) | | 168 (33.5) | | 333 (33.2) |  |
|  | (≤6) | 103 (20.6) | | 85 (16.9) | | 188 (18.7) |  |
| Injury severity based on Glasgow Coma Scale | Median (IQR) | 14 (12-15) | | 14 (11-15) | | 14 (11-15) | .123^t^ |
| Categories of injury severity based on Glasgow Coma Scale (GCS) | (13-15) | 337 (69.9) | | 301 (64.5) | | 638 (67.2) | .175 |
|  | (9-12) | 100 (20.7) | | 119 (25.5) | | 219 (23.1) |  |
|  | (≤8) | 45 (9.3) | | 47 (10.1) | | 92 (9.7) |  |
| Does the severity or multiplicity of injuries exceed local resources and capacity requiring referral? | No | 158 (31.5) | | 154 (30.7) | | 312 (31.1) | .769 |
|  | Yes | 343 (68.5) | | 348 (69.3) | | 691 (68.9) |  |
| Clinical characteristics of participants with neurological injuries | | | | | | | |
| Reported head trauma (impact) irrespective of symptoms | No | 19 (3.8)* | | 35 (7.0)* | | 54 (5.4) | .026* |
|  | Yes | 482 (96.2) | | 467 (93.0) | | 949 (94.6) |  |
| Neurological status | Unresponsive | 5 (1.0) | | 4 (0.8) | | 9 (0.9 | .749 |
|  | Responds to pain | 64 (12.8) | | 63 (12.5) | | 127 (12.7) |  |
|  | Responsive to voice | 123 (24.6) | | 138 (27.5) | | 261 (26.0) |  |
|  | Alert | 309 (61.7) | | 297 (59.2) | | 606 (60.4) |  |
| Symptomatic traumatic brain injury present at admission | No | 154 (30.7) | | 150 (29.9) | | 304 (30.3) | .768 |
|  | Yes | 347 (69.3) | | 352 (70.1) | | 699 (69.7) |  |
| Helmet use | No | | 387 (77.2) | | 362 (72.1) | 749 (74.7) | .062 |
|  | Yes | | 114 (22.8) | | 140 (27.9) | 254 (25.3) |  |
| Basis for suspecting traumatic brain injury | High impact (Yes) | | 43 (8.6) | | 35 (7.0) | 78 (7.8) | .341 |
|  | Loss of consciousness (Yes) | | 286 (57.1) | | 271 (54.0) | 557 (55.5) | .323 |
|  | Post-traumatic convulsions (Yes) | | 21 (4.2) | | 15 (3.0) | 36 (3.6) | .306 |
|  | Post-traumatic amnesia (Yes) | | 26 (5.2) | | 27 (5.4) | 53 (5.3) | .894 |
|  | Post-traumatic headache (Yes) | | 150 (29.9) | | 155 (30.9) | 305 (30.4) | .747 |
|  | Alcohol or drug intoxication (Yes) | | 3 (0.6) | | 2 (0.4) | 5 (0.5) | .652^b^ |
|  | Projectile vomiting (Yes) | | 15 (3.0) | | 7 (1.4) | 22 (2.2) | .084 |
|  | Visible injuries above the clavicle (Yes) | | 237 (47.3) | | 239 (47.6) | 476 (47.5) | .923 |
|  | CSF leak (Yes) | | 46 (9.2) | | 44 (8.8) | 90 (9.0) | .817 |
|  | Focal neurological deficit(s) (Yes) | | 65 (13.0) | | 57 (11.4) | 122 (12.2) | .433 |
| Head and brain CT diagnosis | Epidural hematoma | | 93 (18.6) | | 82 (16.3) | 175 (17.4) | .353 |
|  | Subdural hematoma | | 39 (7.8) | | 56 (11.2) | 95 (9.5) | .068 |
|  | Subarachnoid hematoma | | 24 (4.8) | | 20 (4.0) | 44 (4.4) | .533 |
|  | Intraventricular hemorrhage | | 4 (0.8) | | 5 (1.0) | 9 (0.9) | .740^b^ |
|  | Intraparenchymal hemorrhage | | 16 (3.2) | | 20 (4.0) | 36 (3.6) | .501 |
|  | Traumatic axonal/vascular injury | | 9 (1.8) | | 10 (2.0) | 19 (1.9) | .820 |
|  | Cortical contusions | | 31 (6.2) | | 36 (7.2) | 67 (6.7) | .533 |
|  | Skull fracture | | 57 (11.4) | | 40 (8.0) | 97(9.7) | .068 |
|  | Negative CT results | | 114 (22.8) | | 94 (18.7) | 208 (20.7) | .116 |
|  | Others e.g., maxillofacial | | 12 (2.4) | | 15 (3.0) | 27 (2.7) | .562 |
| Neurosurgical intervention | Watchful waiting | | 370 (73.9) | | 372 (74.1) | 742 (74.0) | .172 |
|  | Craniotomy | | 114 (22.8) | | 122 (24.3) | 236 (23.5) |  |
|  | Decompressive craniectomy | | 17 (3.4) | | 8 (1.6) | 25 (2.5) |  |
| Clinical characteristics of participants with musculoskeletal injuries | | | | | | | |
| Musculoskeletal injury present | No | | 101 (20.2) | | 103 (20.5) | 204 (20.3) | .888 |
|  | Yes | | 400 (79.8) | | 399 (79.5) | 799 (79.7) |  |
| Limb fracture present | No | | 367 (73.3) | | 368 (73.3) | 735 (73.3) | .985 |
|  | Yes | | 134 (26.7) | | 134 (26.7) | 268 (26.7) |  |
| Pelvic fracture | No | | 473 (94.4) | | 468 (93.2) | 941 (93.8 | .436 |
|  | Yes | | 28 (5.6) | | 34 (6.8) | 62 (6.2) |  |
| Tibial fracture | No | | 370 (73.9) | | 374 (74.5) | 744 (74.2) | .814 |
|  | Yes | | 131 (26.1) | | 128 (25.5) | 259 (25.8) |  |
| Femur fracture | No | | 487 (97.2) | | 492 (98.0) | 970 (97.6) | .406 |
|  | Yes | | 14 (2.8) | | 10 (2.0) | 24 (2.4) |  |
| Humerus fracture | No | | 500 (99.8) | | 500 (99.6) | 1000 (99.7) | .564 |
|  | Yes | | 1 (0.2) | | 2 (0.4) | 3 (0.3) |  |
| Radius/ulnar fracture | No | | 496 (99.0) | | 498 (99.2) | 994 (99.1) | .735 |
|  | Yes | | 5 (1.0) | | 4 (0.8) | 9 (0.9) |  |
| Nature of fracture | Open | | 82 (61.2) | | 72 (53.7) | 154 (57.5) | .217 |
|  | Closed | | 52 (38.8) | | 62 (46.3) | 114 (42.5) |  |
| Decision/intent of treatment for the orthopedic or musculoskeletal injury | Conservative ^c^ | | 345 (68.9) | | 363 (72.3) | 708 (70.6) | .231 |
|  | Operative ^d^ | | 156 (31.1) | | 139 (27.7) | 295 (29.5) |  |
| Other associated injuries by body system | Abdomen | | 31 (6.2) | | 24 (4.8) | 55 (5.5) | .328 |
|  | Chest | | 71 (14.2) | | 57 (11.4) | 128 (12.8) | .181 |
|  | Neck | | 41 (8.2) | | 30 (6.0) | 71 (7.1) | .173 |
| Recruitment period | Pre-pandemic | | 82 (16.4) | | 85 (16.9) | 167 (16.7) | .810 |
|  | Post-pandemic | | 419 (83.6) | | 417 (83.1) | 836 (83.4) |  |
| ^a^ Chi-square test of independence except otherwise specified.  ^b^ Fisher’s exact test.  ^t^ Two-sample Wilcoxon rank-sum test.  *Subset of treatment arm whose column proportions differ from each other at *p* < .05 level of statistical significance.  ^c^ Conservative (watchful waiting, physical therapy, casting, braces, splints, pain medication).  ^d^ Operative (open reduction and internal or external fixation or major soft tissue repair requiring theatre).  ^e^ Comorbidity (Diabetes mellitus n=9, Hypertension n=32, HIV/AIDS n=7, Ischemic heart disease n=1, Congestive heart failure n=1, Asthma n=8, Chronic obstructive airway disease n=1, Epilepsy n=3, Malnutrition n=1).  Data are n (%) unless specified otherwise. Denominators for each treatment group are presented in the top row. Missing data were excluded from denominators when computing the percentages. The characteristics of all those included in analyses are demonstrated. | | | | | | | |

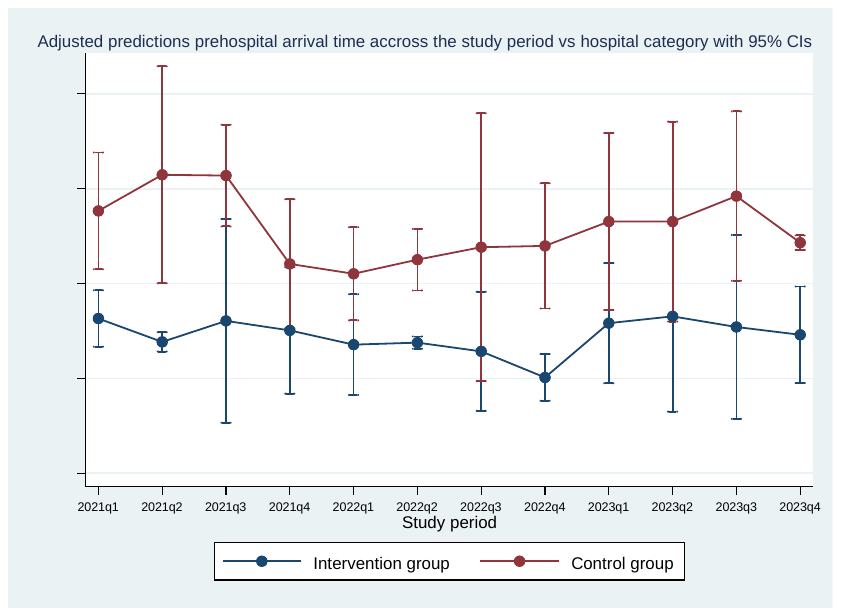

### **Appendix 2: Showing adjusted predictions of prehospital time across study periods.**

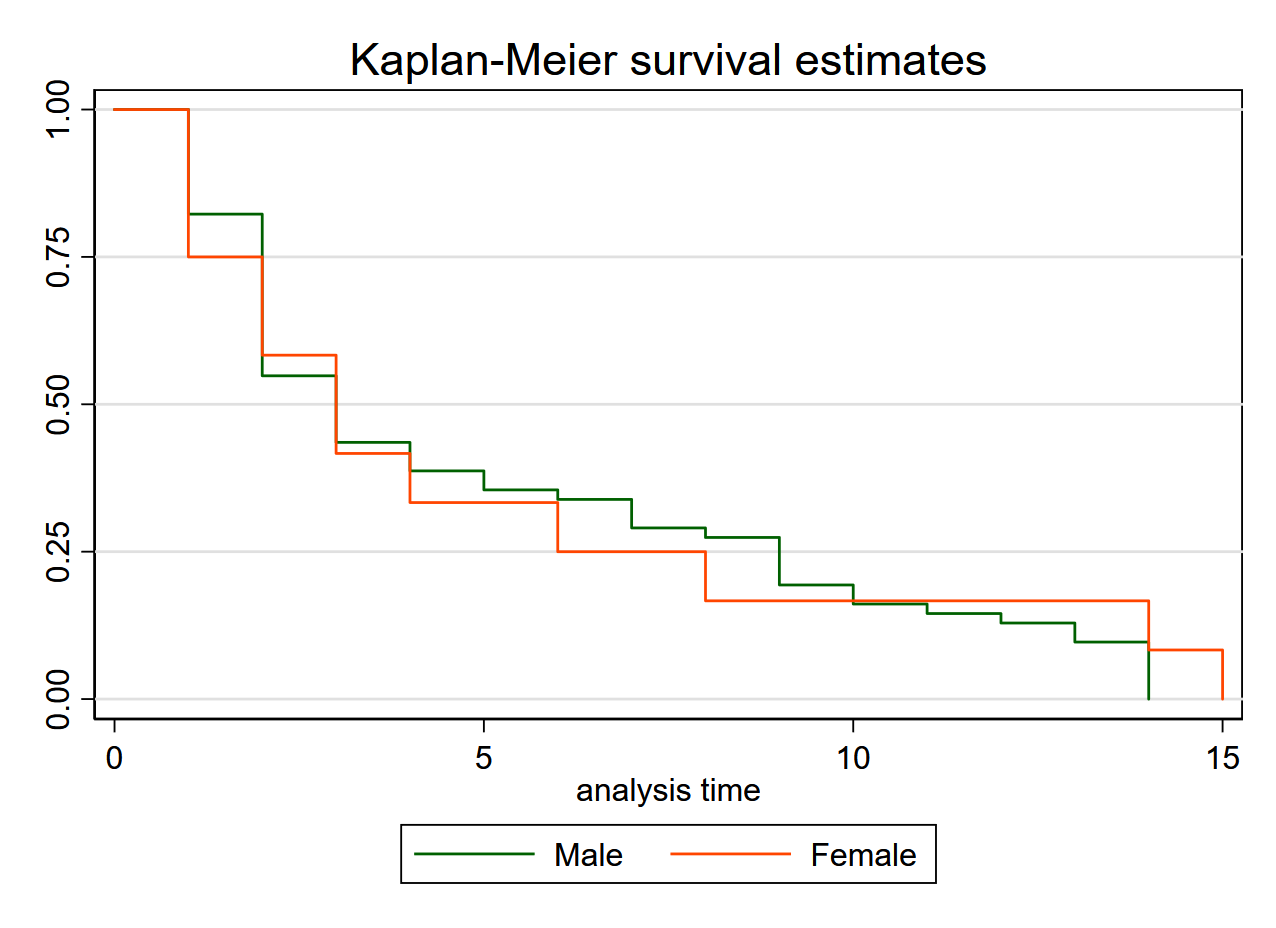

### **Appendix 3: Subgroup analysis of survival time in days stratified by sex.**

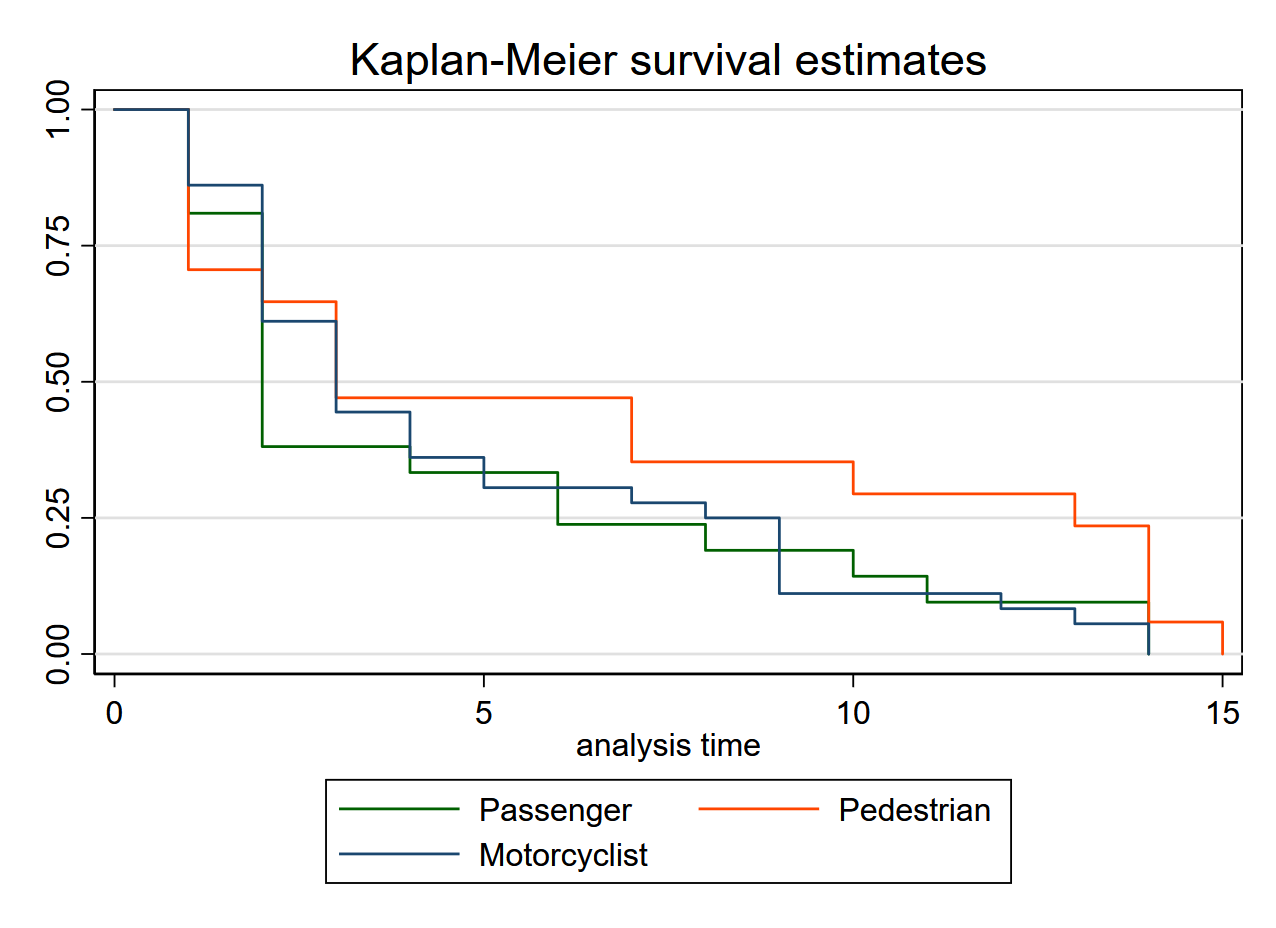

### **Appendix 4: Subgroup analysis of survival time in days by road user category.**

### **Appendix 5: Comparison of Glasgow Outcome Score at 90 days between treatment groups.**

| **GOS** | **Variable** | **Intervention n (%)** | **Control n (%)** | **Overall** |
| --- | --- | --- | --- | --- |
| 1 | Death (clinically confirmed death) | 24 (5.2)* | 58 (13.5)* | 82 (9.2) |
| 2 | Persistent vegetative state (severe damage with prolonged state of unresponsiveness and lack of higher mental function) | 1 (0.2) | 1 (0.2) | 2 (0.2) |
| 3 | Severe disability (severe injury with permanent need for help with daily living) | 17 (3.7) | 26 (6.0) | 43 (4.8) |
| 4 | Moderate disability (no need for assistance in everyday life, employment is possible but may require special equipment) | 123 (26.9) | 101 (23.5) | 224 (25.3) |
| 5 | Good recovery (minimal injury with minor neurological and psychological deficits, patient is independent and employable | 292 (63.9)* | 244 (56.7)* | 536 (60.4) |
|  | Total | 457 | 430 | 887 |
| *Denotes subsets whose column proportions differ significantly from each other at the 95% confidence interval level. | | | | |

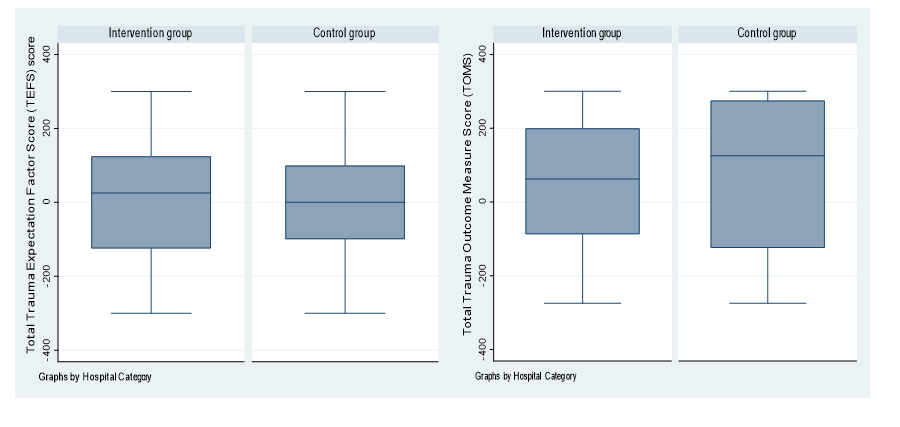

### **Appendix 6: Subgroup analysis of TEFS and TOMS for participants with tibial fractures.**

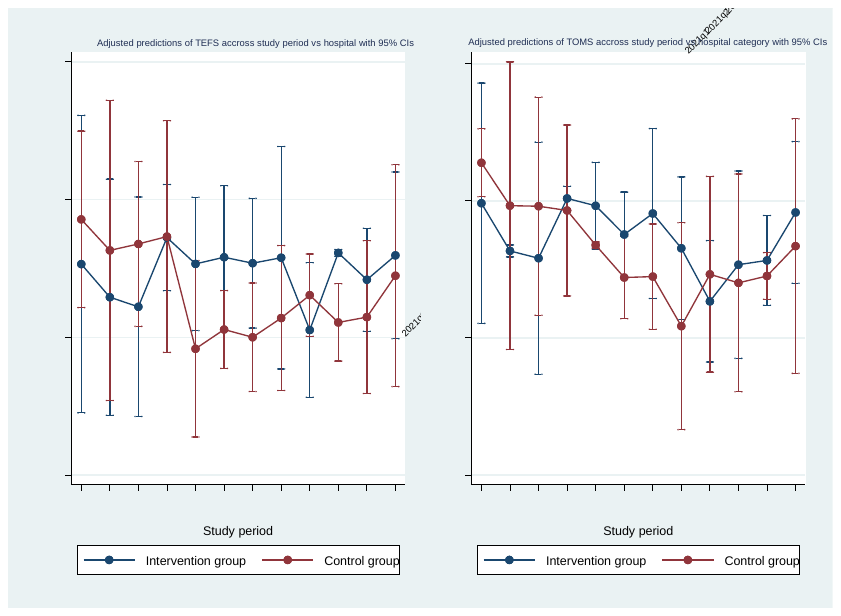

**Appendix 7: Trauma Expectation Factor Scores (TEFS) and Trauma Outcome Measure Scores (TOMS) across study periods.**

### **Appendix 8: Comparison of baseline characteristics amongst those lost to follow-up.**

| **Characteristic** | **Categories** | **Intervention (N=44)**  **n (%)** | **Control (N=72)**  **n (%)** | **Overall (N=116)**  **n (%)** | **p-value** |
| --- | --- | --- | --- | --- | --- |
| Age |  | 28.0 (23.0-40.5) | 30.0 (23-36.5) | 28.0 (23.0-39.0) | .996^c^ |
| Sex | Male | 37 (84.1) | 54 (75.0) | 91 (78.4) | .179^a^ |
|  | Female | 7 (15.9) | 18 (25.0) | 25 (21.5) |  |
| Marital status | Single | 17 (38.6) | 28 (38.9) | 45 (38.8) | .419^a^ |
|  | Married | 24 (54.5) | 41 (56.9) | 65 (56.0) |  |
|  | Divorced | 1 (2.3) | 3 (4.2) | 4 (3.4) |  |
|  | other | 2 (4.5) | 0 (0.0) | 2 (1.7) |  |
| Employment | Formal paid employment | 1 (2.3) | 5 (6.9) | 6 (5.2) | .374^b^ |
|  | Self employed | 68.2 (41.7) | 51 (70.8) | 81 (69.8) |  |
|  | Unemployed | 2 (4.5) | 4 (5.6) | 6 (5.2) |  |
|  | Student | 9 (20.5) | 12 (16.7) | 21 (18.1) |  |
|  | Other | 2 (4.5) | 0 (0.0) | 2 (1.7) |  |
| Commute distance | Median (IQR) | 4 (2.0-17.0) | 5 (3.0-12.5) | 5 (2.7-14.0) | .528^b^ |
| Road user category | Passenger | 11 (25.0) | 32 (44.4) | 43 (37.1) | .055^a^ |
|  | Pedestrian | 9 (20.5) | 16 (22.2) | 25 (21.6) |  |
|  | Motorcyclist | 24 (24.5) | 24 (33.3) | 48 (41.4) |  |
| Mode of arrival | Ambulance | 4 (9.1) | 9 (12.5) | 13 (11.2) | .895^b^ |
|  | Public means (taxi/cycle) | 39 (88.6) | 61 (84.7) | 100 (86.2) |  |
|  | Other | 1 (1.4) | 2 (2.8) | 3 (2.6) |  |
| Prehospital care (first aid) received? | Yes | 26 (59.0) | 40 (55.6) | 66 (56.9) | .709^a^ |
|  | No | 18 (40.9) | 32 (44.4) | 50 (43.1) |  |
| Who gave first aid? | Health worker | 14 (31.8) | 24 (33.3) | 38 (32.8) | .825^b^ |
|  | Lay bystander/police | 4 (9.1) | 8 (11.1) | 12 (10.3) |  |
| Mechanism of injury | Motorcycle-motorcycle crash | 19 (43.2) | 41 (56.9) | 60 (51.7) | .357^b^ |
|  | Motorcycle-pedestrian | 13 (29.5) | 20 (27.8) | 33 (28.4) |  |
|  | Motorcycle-car crash | 9 (20.5) | 9 (12.5) | 18 (15.5) |  |
|  | Motorcycle static object | 3 (6.8) | 2 (2.8) | 5 (4.3) |  |
| Comorbidity (chronic medical illness present) | Hypertension | 3 (100.0) | 1 (33.3) | 4 (66.7) | .400^b^ |
|  | Asthma | 0 (0.0) | 2 (66.7) | 2 (33.3) |  |
| Injury severity (KTS) | Mild (9-10) | 25 (56.8) | 41 (56.9) | 66 (56.9) | .682^b^ |
|  | Moderate (7-8) | 14 (31.8) | 26 (36.1) | 40 (34.5) |  |
|  | Severe ≤ 6 | 5 (11.4) | 5 (6.9) | 10 (8.6) |  |
| Score for serious injuries | > one | 4 (9.1) | 8 (11.1) | 12 (10.3) | .661^b^ |
|  | one | 32 (72.7) | 52 (72.2) | 87 (75.00 |  |
|  | None | 5 (11.4) | 12 (16.7) | 17 (14.7) |  |
| **^a^** Chi-square test for independence, ^b^ Fisher’s Exact test; ^c^ Two-sample Wilcoxon rank sum test | | | | | |

### **Appendix 9: Reflexivity statement regarding collaborative partnerships. [1]**

| **Item** | **Question** | **Answer** |
| --- | --- | --- |
| Study conceptualization | How does this study address local research and policy priorities? | This study addressed rural trauma training and coordination which are critical issues in Uganda due to its high injury burden and inadequate human resources for health. |
|  | How were local researchers involved in the study? | The PI was a local researcher who leveraged locally available expertise to offer the rural trauma team development training intervention. The local research team assisted with necessary ethical, logistical, and administrative clearances. |
| Research management | How has funding been used to support the local research team? | HIC partners hosted LMIC (PI) through a funded doctoral researcher position at the University of Turku (Finland) to develop local research capacity. The Centre for Health Equity in Surgery and Anesthesia (CHESA) at University of California San Francisco (USA) provided a travel grant to PI to facilitate dissemination results through participation in CHESA fellowship. |
| Data acquisition and analysis | How are research staff who conducted data collection acknowledged? | Research staff who met the authorship criteria detailed in the international committee of medical journal editors (ICMJE) guidelines were included as authors whereas those who did not meet these criteria but made substantial contributions are acknowledged in the acknowledgement section. |
|  | How have members of the research partnership been provided with access to the study data? | The data collection team were issued login access to the raw data in REDCap and the final dataset is accessible to all partnership through unrestricted access publishing. |
|  | How were data used to develop analytical skills within the partnership? | LMIC PI developed a locally contextualized data collection tool. HIC partners provided working space and software to LMIC PI to develop capacity for data analysis. |
| Data interpretation | How have research partners collaborated in interpreting the data? | LMIC PI analysed and interpreted the data with visualization and cross-validation support from an internal and external biotechnicians from HICs. |
| Drafting and revising for intellectual content | How were research partners supported to develop writing skills? | LMIC PI was supported to attend academic writing courses at the University of Turku in Finland. |
|  | How will research products be shared to address local needs? | The study findings were accepted in form of abstract for oral presentation to stakeholders under the theme “equitable and sustainable strategies for injury and violence prevention) during WHO safety 2024 (world conference on injury prevention and safety promotion 2-4 September 2024 in New Delhi, India) and a local regional dissemination is planned at the 24^th^ scientific conference for the College of Surgeons of East, Central and Southern Africa during 2-6 December 2024 in Harare, Zimbabwe. |
| Authorship | How is the leadership, contribution, and ownership of this work by LMIC researchers recognized within the authorship? | Authorship was based on the ICMJE criteria. LMIC PI is the first author and HIC supervisors are the last authors. |
|  | How have early career researchers across partnership been included within the authorship teem? | Early career researchers whose contributions met the inclusion criteria were included as authors. |
|  | How has gender balance been addressed within the authorship? | The authorship was based on ICJME guidelines as per trial protocol without gender bias. |
| Training | How has the project contributed to training LMIC researchers? | The project is part of the LMIC PI’s body of work for a doctoral degree. |
| Infrastructure | How has project contributed to improvements in local infrastructure? | We trained a total of five hundred rural trauma care frontliners during this project to build capacity of injury care in LMICs. Further, we piloted a motorcycle trauma outcome (MOTOR) registry in parallel to the trial. |
| Governance | What safeguarding procedures were used to protect local study participants and researchers? | The study was conducted in accordance with the Uganda National Council for Science and Technology guidelines on human subjects as research participants. |
| [1] B. Morton *et al.*, “Consensus statement on measures to promote equitable authorship in the publication of research from international partnerships.,” *Anaesthesia*, vol. 77, no. 3, pp. 264–276, Mar. 2022, doi: 10.1111/anae.15597. | | |
